# Exposure to per- and polyfluoroalkyl substances associates with altered lipid profile of breast milk

**DOI:** 10.1101/2021.02.10.21251515

**Authors:** Santosh Lamichhane, Heli Siljander, Daniel Duberg, Jarno Honkanen, Suvi M. Virtanen, Matej Orešič, Mikael Knip, Tuulia Hyötyläinen

**Author notes:** Shared senior authorship (Matej Orešič, Mikael Knip, Tuulia Hyötyläinen). The authors declare that they have no competing interests.

## Abstract

**Background:** Chemical composition of human breast milk is highly variable inter- and intra-individually. Environmental factors are suspected to partly explain the compositional variation, however, their impact on breast milk composition is currently poorly understood.

**Objectives:** We sought (1) to define the impact of maternal exposure to per- and polyfluoroalkyl substances (PFAS) on lipid composition of human breast milk, and (2) to study the combined impact of maternal PFAS exposure and breast milk lipid composition on the growth of the infants.

**Methods:** In a mother-infant study (n=44) we measured the levels of PFAS and lipids in maternal serum and conducted lipidomics analysis of breast milk at birth and at 3 months of infant age, by using ultra high performance liquid chromatography combined with quadrupole-time-of-flight mass spectrometry. Maternal diet was studied by a validated food frequency questionnaire.

**Results:** PFAS levels were inversely associated with total lipid levels in the breast milk collected at birth. In the high exposure group, the ratio of acylated saturated and polyunsaturated fatty acids in triacylglycerols was increased. Moreover, high exposure to PFAS associated with the altered phospholipid composition, which was indicative of unfavorable increase in the size of milk fat globules. These changes in the milk lipid composition were further associated with slower infant growth and with elevated intestinal inflammatory markers.

**Discussion:** Our data suggest that the maternal exposure to PFAS impacts the nutritional quality of the breast milk, which, in turn, may have detrimental impact on the health and growth of the children later in life.

## Introduction

Breast milk (BM) is considered as an optimal source of nutrition for infants for the first 6 months of life, a crucial period for immune system development, metabolic and endocrine programming for growth, development, and lifelong health (*1*). Thus, the composition of the BM is essential for neonatal health outcomes and the lipid composition has been associated with adequate growth, neurocognitive development and function, regulation of inflammation and infection risk, and risk of later metabolic and cardiovascular disease in adulthood (*2*). Particularly, the fatty acid composition of the lipid part of the BM has been linked to the development of gut microbiota.

The chemical composition of human milk changes from colostrum to late lactation, and varies within feeds, diurnally, and between mothers (*1*), being influenced by both maternal and environmental factors and showing high inter- and intra-individual variability (*3*). It has been shown that birth weight, gestational age, infant age/stage of lactation has an impact on the composition, but less is known how the maternal factors such as lifestyle (except for the mother’s diet) and socio-demographic factors are affecting the BM composition. Interestingly, the socioeconomic status and geographical location were found to have an impact particularly on the lipid composition of the milk (*3*). For example, a more favourable balance between omega-3 and omega-6 polyunsaturated fatty acids (PUFAs) was observed in BM of Swedish women compared with Chinese women (*4*). Differences in diet may explain some of the findings, however, it is likely that there are also other underlying factors that drives the differences in BM composition. One of these factors may be the environmental toxins that the mother has been exposed to, mainly *via* her diet.

One specific class of chemicals that the humans are widely exposed to include per- and polyfluoroalkyl substances (PFAS), which are a class of anthropogenic substances that have been manufactured for over six decades. They have been used in a wide range of commercial processes and consumer products and due to their persistent nature, they are widely distributed in the environment, including water, soils, sediment, wildlife and humans (*5*). Interestingly, several epidemiological studies have reported that maternal PFAS levels are inversely associated with the duration of breastfeeding, even after controlling for potential confounding factors such as prior breastfeeding, smoking, alcohol intake, body mass index (BMI), education and primiparity (*6–8*). However, the physiological reasons for insufficient milk production are currently poorly understood. The main research focus has been hormonal abnormalities, maternal disease and contraindications. In animal studies, gestational exposure to PFAS was found to reduce mammary differentiation in the exposed dams and to stunted mammary epithelial branching and development in the pups (*9*). Furthermore, perfluorooctanoic acid (PFOA) stimulates mammary gland development by promoting steroid hormone production in ovaries and increasing the levels of a number of growth factors in mammary glands (*10*), and exerts its effect *in vivo* through estrogen receptor (ER) dependent mechanisms (*11*). Possible modes of action by which PFAS interfere with successful acini maturation could include modulation of cell-extracellular matrix (ECM) interactions, cell–cell adhesion and/or intercellular communication, and their combinations (*9*). Moreover, the exposure to PFOA and perfluorooctanesulfonic acid (PFOS) was found associated with prolactin levels (*12, 13*). It is thus conceivable that accumulated PFAS exposures affect the ability to lactate. Exposures to several other environmental toxicants, including 2,3,7,8-tetrachlorodibenzo-p-dioxin (TCDD), were also found implicated in impaired mammary differentiation and lactation (*14, 15*).

Since multiple studies show that the exposure does impact the mammary gland differentiation, it is plausible that disruption of this complex process leads to changes in the BM composition. Currently, there are very limited studies on the impacts of environmental exposures on the molecular lipid content of BM. Recent study in rats showed that exposure to bisphenol A (BPA) induces a delay in the functional differentiation of the mammary gland during secretory activation, resulting also in an alteration of the lipid profiles in the milk (*16*). However, no previous studies have specifically investigated the impact of PFAS exposure on the lipid composition of BM. It has been suggested that milk lipid formation involves sequentially regulated lipogenic, transport and secretion patterns that are functionally linked to the maturation of alveoli into milk-secreting structure (*13*). Another regulator of lipid metabolism in the mammary gland is the peroxisome proliferator-activated receptor gamma (PPARγ) (*17*), which plays a role in controlling milk fat synthesis, either directly or via the activation of the transcription regulators sterol regulatory-element binding protein 1 (SREBP1) and liver X receptor alpha (LXR-alpha).

Herein, we investigate how the PFAS levels in maternal blood are associated with the lipidomic profile of BM. We also study how these lipid changes in BM associate with infant growth.

## Methods

### Regulatory and ethics status

The study followed the guidelines of the Declaration of Helsinki for research on human participants. The study was carried out according to good clinical practice. The study protocol was approved by the Joint Municipal Authority of the Pirkanmaa Hospital District, Finland (no. R11166). The parents gave their written informed consent, which was overseen by the Ethical Committee of the Joint Municipal Authority of the Pirkanmaa Hospital District. The study is registered under Clinicaltrials.gov Identifier NCT01735123.

### Study population

Pregnant Finnish women were recruited from January 28, 2013 to February 26, 2015, in the context of the EDIA (Early Dietary Intervention and Later Signs of Beta-Cell Autoimmunity: Potential Mechanisms) study, which is a small-scale intervention trial comparing weaning infants onto an extensively-hydrolyzed milk formula vs. a conventional cow’s milk-based formula. Families were contacted at the time of the fetal ultrasonography visit, which is arranged for all pregnant women in Finland around gestational week 20. Written, informed consent was signed by the parents to permit analysis of their HLA genotype to exclude infants without HLA-conferred susceptibility to T1D. Patient consent was overseen by the Ethical Committee of the Joint Municipal Authority of the Pirkanmaa Hospital District. At this point, 68% of the infants to be born were excluded. Separate informed consent was obtained from eligible parents at the beginning of the third trimester to analyze the offspring’s genotype and to continue in the intervention study. The cord blood from 309 newborn infants was screened to determine the HLA genotype, as previously described (*18*). The degree of HLA susceptibility to T1D was divided into six categories (high-risk, moderate-risk, low-risk, neutral, protective and strongly protective genotypes), as earlier defined (*19*). Eighty-seven offspring were eligible, as they belonged to the three highest HLA risk categories. Out of the eligible infants 73 joined the intervention trial. A serum sample and a BM sample was collected from the participating mothers at two time points for the current study; in the delivery hospital (2-4 days after delivery) and when the baby was 3-month-old. In the current study we included 44 mothers (**Table 1)** with two serum and two BM samples available and with their diet during pregnancy assessed with a validated, semiquantitative food frequency questionnaire (FFQ) (*20*). Food and individual nutrient intakes were calculated using the national food composition database Fineli (https://fineli.fi/fineli/en/index), using the in-house software (Finessi) of the Finnish Institute for Health and Welfare, Finland (*21*).

**Table 1.** Demographic characteristics of the study population.

| **Parameter** | **Median (range)** |
| --- | --- |
| **Maternal delivery age (years)** | 32.47 (24.89-45.80) |
| **Gestational age (weeks)** | 40.28 (36.57-42.42) |
| **Post-pregnancy BMI (kg/m2)\*** | 25.46 (18.49-33.7) |
| **Mother height (cm)** | 166.5 (156-178) |
| **Child weight (kg)\*\*** |  |
| 0 months (birth) | 3.55 (4.37-2.65) |
| 3 months | 6.44 (4.33-8.12) |
| 6 months | 7.9 (6.31-10.60) |
| 9 months | 9.06 (7.23-11.78) |
| 12 months | 9.90 (8.06-13.22) |
| **Child length (cm)\*\*** |  |
| 0 months (birth) | 50.5 (46.0-54.0) |
| 3 months | 62.2 (56.2-65.20) |
| 6 months | 68.0 (63.0-71.4) |
| 9 months | 72.7 (67.9-76.9) |
| 12 months | 76.4 (71.0-81.7) |
| **Sex of the baby (male/female)\*\*** | 20/19 |
| **Fecal beta-defensin-2 (ng/ml)\*\*** |  |
| 3 months | 288 (61- 1494) |
| 6 months | 96 (18-1017.1) |
| 9 months | 38 (10-642) |
| 12 months | 44 (0-332) |
| **Faecal calprotectin (mg/kg)\*\*** |  |
| 3 months | 241.9 (61.6 - 925.8) |
| 6 months | 46.8 (421-4.5) |
| 9 months | 16.4 (1.1-205) |
| 12 months | 14 (0-89.3) |

\*For a subgroup of 42 subjects.
\*\*For a subgroup of 39 subjects.

### Analysis of molecular lipids

#### Sample extraction

30 µl BM sample or 10 µl of serum was extracted with using a modified version of the previously published Folch procedure (*22*). In short, 10 µL of 0.9% NaCl and, 120 µL of CHCl3: MeOH (2:1, v/v) containing the internal standards (c = 2.5 µg/mL) was added to the sample. The standard solution contained the following compounds: 1,2-diheptadecanoyl-sn-glycero-3-phosphoethanolamine (PE(17:0/17:0)), N-heptadecanoyl-D-erythro-sphingosylphosphorylcholine (SM(d18:1/17:0)), N-heptadecanoyl-D-erythro-sphingosine (Cer(d18:1/17:0)), 1,2-diheptadecanoyl-sn-glycero-3-phosphocholine (PC(17:0/17:0)), 1-heptadecanoyl-2-hydroxy-sn-glycero-3-phosphocholine (LPC(17:0)) and 1-palmitoyl-d31-2-oleoyl-sn-glycero-3-phosphocholine (PC(16:0/d31/18:1)), were purchased from Avanti Polar Lipids, Inc. (Alabaster, AL, USA), and, triheptadecanoylglycerol (TG(17:0/17:0/17:0)) was purchased from Larodan AB (Solna, Sweden). The samples were vortex mixed and incubated on ice for 30 min after which they were centrifuged (9400 × g, 3 min). 60 µL from the lower layer of each sample was then transferred to a glass vial with an insert and 60 µL of CHCl3: MeOH (2:1, v/v) was added to each sample. The samples were stored at −80 °C until analysis.

#### Instrumental analysis

The lipidomic analyses were done using an ultra-high-performance liquid chromatography quadrupole time-of-flight mass spectrometry method (UHPLC-Q-TOF-MS from Agilent Technologies (Santa Clara, CA, USA). The analysis was done on an ACQUITY UPLC® BEH C18 column (2.1 mm × 100 mm, particle size 1.7 µm) by Waters (Milford, USA). Internal standard mixture was used for normalization and lipid-class specific calibration was used for quantitation as previously described (*23*). MS data processing was performed using open source software MZmine 2.52 (*24*).

### PFAS analyses

#### Sample extraction

The serum samples were extracted as follows after randomization of the samples. 90 µl of acetonitrile (containing the BA+PFAS internal standard mixture; c=200 ng/mL PFASs and 1000 ng/mL Bile acids in acetonitrile) was added to the 40 µl of serum samples. The samples were then vortexted and centrifuged (9600 RCF, 10 minutes). 90 µl of the supernatant was collected and evaporated to dryness. The samples were reconstituted with 40 µl of MeOH/H2O (40%/60%).

#### Instrumental analysis

PFASs were analysed on a UHPLC-qTOF/MS (Agilent Technologies, Santa Clara, CA, USA) with Acquity UPLC®, BEH C18 (2.1 x 100 mm, particle size 1.7 µm) (Waters Corporation, Milford, MA, USA) column at 50°C with a C18 pre-column for column protection (Waters Corporation, Wexford, Ireland). Mobile phases used for the sample analysis were A: 2mM NH4Ac in H2O:MeOH (70:30) and B: 2mM NH4Ac in MeOH. NH4Ac was used as ionization agent. The samples were kept at 10°C during the whole sample acquisition and 1 µl of the sample volume was injected. The flow rate was set to 0.4 ml/min and the gradient started with 95%A and 5%B with a change after 1.5 minute to 70%A and 30%B, which followed a change after 4.5 minutes to 30%A and 70%B, the last change was after 7.5 minutes with 100%B until the end of run. Gradient program was 18 minutes long. 13 minutes for sample analysis and 5 minutes for clean-up, including equilibration in the end of the run. Maximum pressure limit in the binary pump was set on 850 bar. Dual jet stream electrospray (dual ESI) ion source was used and the ion polarity was on negative mode. The capillary voltage and the nozzle voltage were kept at 4500 V and 1500 V. The N2 pressure was set on 21 psi, with the sheath gas flow as 11 L/min and temperature at 379°C for the nebulizer. The data was acquired with MassHunter B.06.01 software (Agilent Technologies, Santa Clara, CA, USA). MS data processing was performed using open source software MZmine 2.52 (*24*).

### Data pre-processing

Lipids were processed separately from BA and PFAS data. Mass spectrometry data was pre-processed with MZmine 2.53 (Pluskal et al., 2010). Peak detection with a noise level of 1000 was performed first, following with ADAP chromatogram builder including group intensity threshold to be as noise level (1000), minimum highest intensity 10 and m/z tolerance 0.009 m/z or 8 ppm. Next, chromatogram deconvolution was performed with local minimum search as algorithm with a 70% chromatographic threshold, 0.05 minutes of minimum retention time range, 5% minimum relative height, 2250 minimum absolute height, 1 as minimum ratio of peak top/edge and peak duration range in minutes from 0.08 to 5.00. Isotopic peak grouper was done next with m/z tolerance of 0.05 m/z or 5 ppm, tR tolerance was set on 0.05 minutes and a maximum charge of 2. For the alignment of peak lists, a Join alignment algorithm was performed with m/z tolerance as 0.006 or 10.0 ppm and a weight of m/z as 2 with a tR tolerance of 0.1 and a weight of tR 1. Filtering with feature list rows filter was done next with three steps. First step rows that match with all criteria were kept with a retention time range from 2 to 12 and m/z of 369-1200. Second filtering step removed rows that match with all criteria with a tR range of 2-4 and m/z of 800-1200. Third filtering step removed rows that match with criteria with a tR range of 4-8 and m/z of 370-500. Next, gap filling with peak finder was done with m/z tolerance of 0.006 m/z or 10.0 ppm, tR tolerance of 0.1 minutes and with intensity tolerance as 50%. Last, the identification with a custom data base was done to identify the peak list with compound names.

### Quantitation and quality control

Quantification of lipids was performed using a 7-point internal calibration curve ranging from 0.1 up to 5 µg/mL. Quality control was performed throughout the dataset both for lipidomics and PFAS analysis by including blanks, pure standard samples, extracted standard samples and control plasma samples. Relative standard deviations (%RSDs) for lipids in the pooled samples (*n* = 6 and 7 for breastmilk and serum, respectively) were on average 15.9 % and 11.8% for BM and serum lipids, respectively. For PFAS, RSD concentrations in the pooled serum samples (*n* = 7) was on average 21.8 %.

### Analysis of fecal calprotectin and HBD-2

Stool samples from the infants were collected and frozen immediately at home (−20°C). Parents brought the frozen samples to the study center, after which samples were stored at −70°C until analyzed. Fecal calprotectin and HBD-2 levels were analyzed with commercial ELISA kits according to manufacturer’s instructions (Calpro AS, Lysaker, Norway, and β-Defensin 2 ELISA Kit, Immundiagnostik, Bensheim, Germany) (*25, 26*). Briefly, approximately 100 mg of feces was obtained from each frozen sample. Extraction buffer was then added at a dilution of 1:50 for both HBD-2 and calprotectin. Fecal material with the extraction buffer was vortexed for 30 seconds and mixing was continued in a shaker at 1000 r.p.m. for 3 minutes or until solid particles had dissolved. Samples were then centrifuged for 10 min at 10 000 g at room temperature and the supernatants were collected and stored at −20°C until measured.

### Statistical analysis

All statistical analyses were on individual lipid intensity level and lipid class specific analysis. The individual lipid in the lipidomics dataset were clustered to represent a specific lipid class. Individual lipid concentrations in each lipid class were first median normalized, summed, then subsequent data analysis considering each lipid class as a variable was performed. To identify any lipid/lipid class changing in value across various levels of PFAS exposure, the maternal exposure level were first assigned to four quartiles of total PFAS exposure, defined as the sum of their raw PFAS exposures. Subsequently, Wilcoxon rank-sum test was used to compare lipid profile between the higher (Q4) and the lower (Q1) exposure level on the BM; samples (at delivery or at 3 months). Wilcoxon rank-sum test was performed in MATLAB 2017b (MathWorks Inc., Natick, MA) using Statistical Toolbox. p-values < 0.05 (two-tailed) were considered significant. To subsequently visualize the lipids levels as violin plot the ggplot2 R package was used. The fold difference was calculated by dividing the mean concentration of a lipid species in one group by another, for instance mean concentration in the the higher exposure level (Q4) by the mean concentration in lower exposure level (Q1) and then illustrated by bargraph using the ggpubr R package (version 0.4). Spearman correlation coefficients were calculated using the Statistical Toolbox in MATLAB 2017b and p-values < 0.05 (two-tailed) were considered significant for the correlations. The individual spearman correlation coefficients (R) were illustrated as a heatmap using the “corrplot” package (version 0.84) for the R statistical programming language (https://www.r-project.org/).

## Results

### Maternal PFAS levels and breast milk lipidome

Fourty-four mothers were included in the study (**Table 1**), with breast milk samples at two time points (at delivery, 0 months, and after 3 months). Maternal PFAS levels were measured in serum samples collected 3 months after the delivery. Six PFAS were detected and quantified from the samples, namely PFDA, PFHxS, PFNA, PFOA and two isomers of PFOS (linear and branched) (**Table 2**). Based on the total PFAS concentration, the group was divided into four exposure quartiles. In addition, lipidomic profiles of BM and blood were measured at both time points. 261 lipids were identified from the samples, with triacylglycerols (TG) being the most abundant group. The lipids identified comprised mainly of ceramides (Cer), including hexosylceramides (HexCer) and dihexosylceramides (diHexCer), cholesterol esters (CE), diacylglycerols (DG), lysophosphatidylcholines (LPC), phosphatidylcholines (PC), phosphatidylethanolamines (PE), phosphatidylinositols (PI), and sphingomyelins (SM). The levels of several Cer and PEs were higher at the 3-month time point as compared to 0-months, while CEs, PCs, HexCer and TGs with polyunsaturated fatty acyls were significantly lower at the later time point. The BM lipids were not significantly associated with the number of previous births, maternal age or BMI, except for a weak positive correlation between birth order and HexCer, diHexCer and PEs (**Supplementary Figure 1**).

**Table 2.** Concentration of PFAS measured in the serum of pregnant mothers.

|  | **Concentration (ng/ml)**<br>**Median (Min., Max.)** |
| --- | --- |
| **PFDA** | 0.61 (0.20, 1.00) |
| **PFHxS** | 0.23 (0.15, 0.38) |
| **PFNA** | 1.58 (0.24, 5.31) |
| **PFOA** | 4.74 (1.22, 11.33) |
| **Br-PFOS** | 5.93 (2.04, 15.12) |
| **L-PFOS** | 7.24 (3.02, 19.48) |

### Impact of PFAS exposure on the breast milk lipidome

The amount of total lipids had a significant inverse correlation with the total PFAS levels (sum of concentrations of all PFAS) in the BM samples collected at the time of delivery (**Figure1A**), with a fold change of −1.7 between the highest and lowest quartiles (p= 0.0052), while at the later, 3-month time point, there were no correlations between the total lipids and PFAS levels. At the level of individual lipids, at birth, 49 lipids were different between the highest and lowest quartiles (**Figure 1B, Supplementary Table 1**), while at the 3-month time point, only a few lipids showed significant changes. Overall, lipids containing saturated fatty acyls were increased in the highly exposed mothers, while CEs and lipids containing unsaturated fatty acyls were decreased.

**Figure 1.**
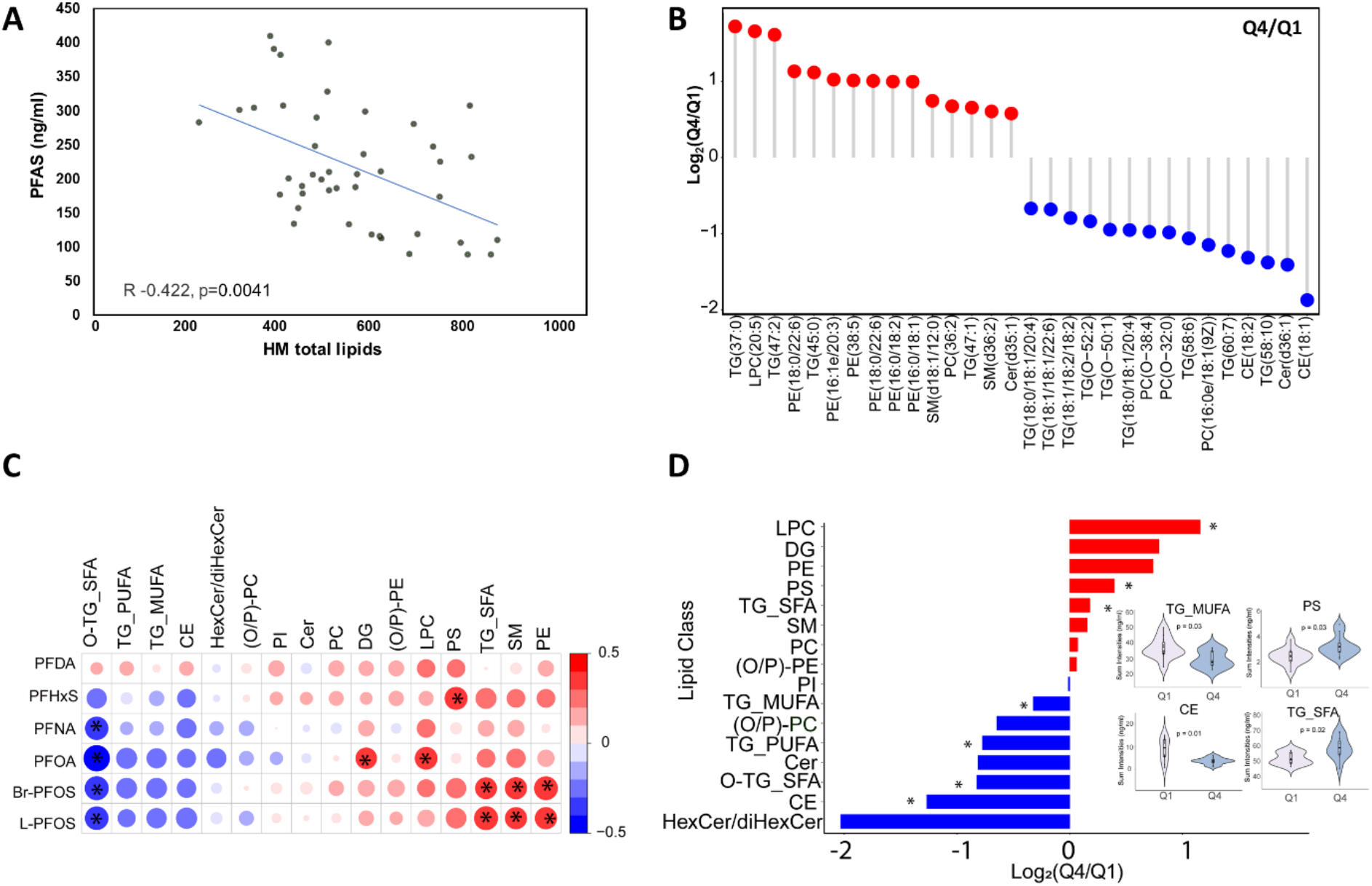
Impact of exposure on BM lipid levels. (**A**) Association of total PFAS and total lipids. (**B**) Significantly different lipids for total PFAS quartile 4 (Q4, high exposure) *vs*. quartile 1 (Q1, low exposure). (**C**) Correlation plot of PFAS exposure and BM lipid classes. Positive correlations in red, inverse correlations in blue. Dot size for each pairwise correlation corresponds to the strength of the calculated correlation. (**D**) Overall difference in lipids classes for total PFAS quartile 4 (Q4, high exposure) *vs*. quartile 1 (Q1, low exposure). *p < 0.05.

At the lipid class level, significant differences between the high and low exposure groups were observed in the BM levels of LPCs, PCs, and TGs in BM samples collected at birth. Concentrations of LPCs and TG_SFA (TGs enriched with fatty acyls of low saturation level, *i*.*e*., 0-2 double bonds) were increased, while TGs enriched with polyunsaturated fatty acyls (TG_PUFA) were diminished in the high exposure group (**Figure 1C-D**). The ratio between TG_SFA and TG_PUFA was increased due to the exposure (**Figure 2A**). The lipids being upregulated in high PFAS exposure group were mainly those lipids that contained saturated or monounsaturated fatty acyls, while those being downregulated were mainly those with polyunsaturated fatty acyls, including docosahexaenoic (DHA) (**Figure 2A**). Also the ratio of PC to PE as well as the TG ratio of PUFA/SFA were significantly downregulated in the high exposure group (**Figure 2B**). At 3-month time point, fewer differences were observed in BM, with only DGs and CEs being different between the high- and low-exposure groups.

**Figure 2.**
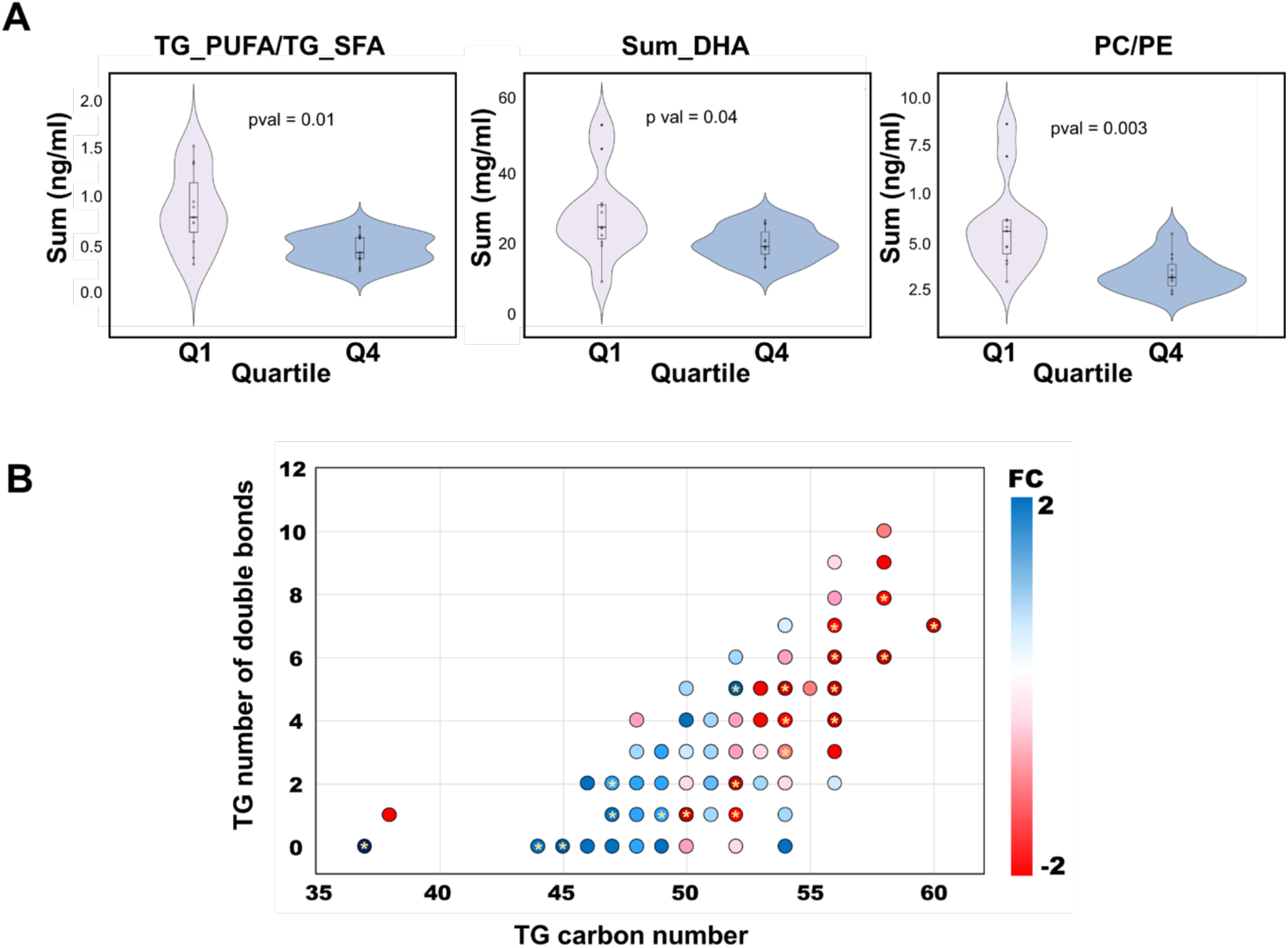
Impact of high and low PFAS exposure on structurally specific BM lipids. (**A**) Comparison of lipid concentration (potential docosahexaenoic (PDHA), PC /PE and PUFA/SFA) between high exposure (Q4) and low exposure (Q1). (**B**) Differences in individual TGs between PFAS quartile 4 (Q4, high exposure) *vs*. quartile 1 (Q1, low exposure). The x-axis is the acyl carbon number and the y-axis is the acyl double-bond count.

### Association of infant growth and BM lipidome

Infant growth data were available at 3, 6, 9 and 12 months of age, alongside the data on gut inflammatory markers (fecal calprotectin and fecal beta2 defensin). PFAS exposure had a trend for inverse associations with the child growth (weight) at 3 months of age, and the measurement of length (linear growth) showed weak associations, not reaching statistical significance. However, the PE, PC, (O/P)-PE, PS, and SM in BM had an inverse association with the infant growth rate (weight gain) at 3 months of age, and the PE and PC also at 6 months (**Figure 3**). Also the PC/PE level was positively associated with early growth (weight). At the level of individual molecular lipids, 77 lipids had a significant association with the offspring growth [weight or length at at least one time point (**Supplementary Figure 2**)]. Fecal calprotectin showed a significant, positive association with several PFAS and specific BM lipid classes.

**Figure 3.**
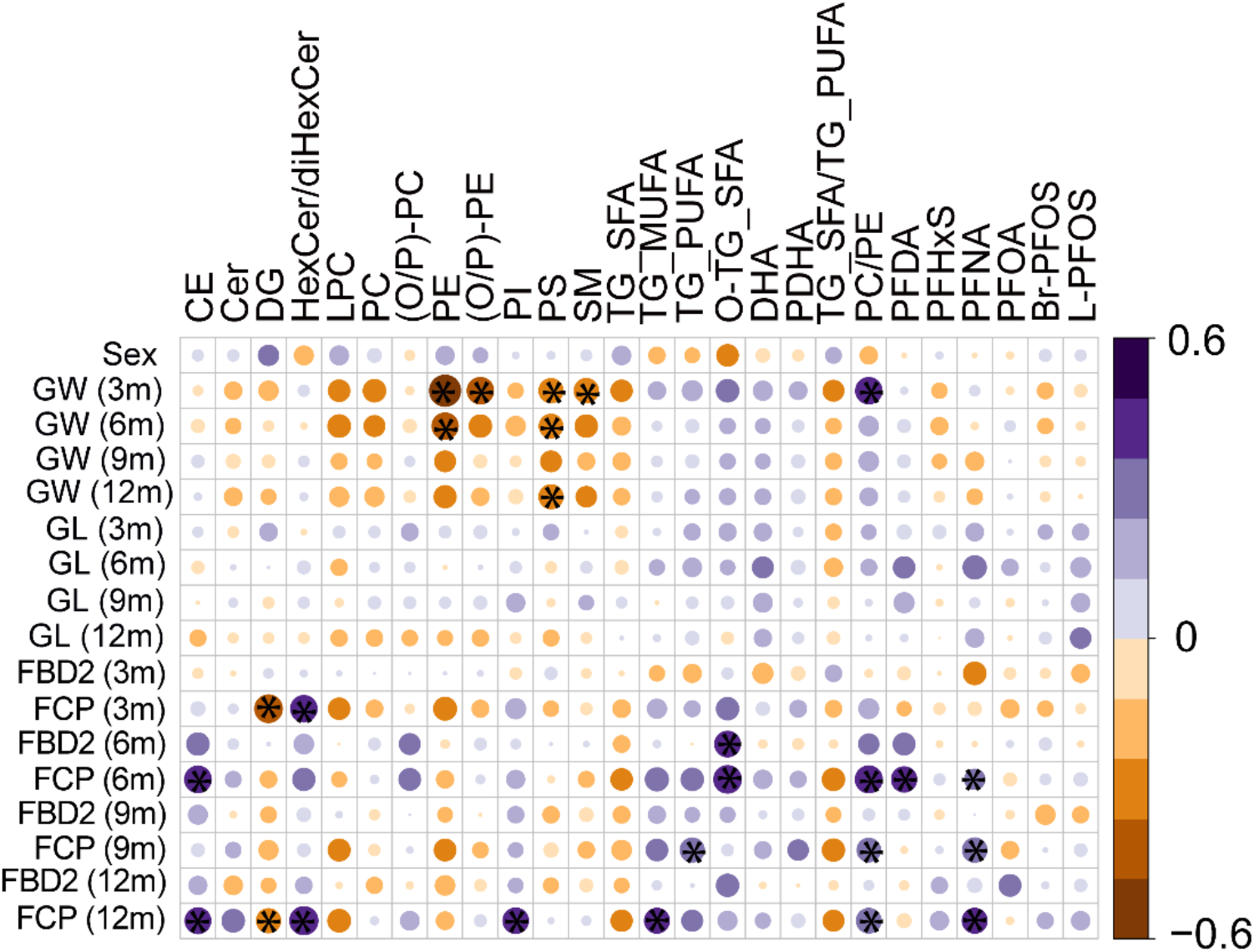
Associations of BM lipids and PFAS with growth and markers of intestinal inflammation in the offspring.Pairwise Spearman correlation as calculated between BM lipids and PFAS with offspring weight and markers of gut inflammation. Here, GW is growth in weight (kg/year), GL is growth in length (cm/year), FBD2 is Fecal beta defensin 2, FCP is fecal calprotectin. Correlations were calculated between simultaneous measurement at 3, 6, 9, and 12 months of age. GL and GW were calculated based on length in cm (or weight in kg) at given months – birth length in cm (or weight in kg))/time (measurement age (in years) at the given sampling day) respectively. Positive correlations in purple, inverse correlations in orange. Dot size for each pairwise correlation corresponds to the strength of the calculated correlation. *p < 0.05.

### Impact of maternal diet on the breast milk lipidome

The impact of maternal food consumption during pregnancy on the lipid profiles in BM was investigated using correlation analysis and partial correlation network analysis. In the paired serum-BM samples, the lipids in serum had a very low association with BM lipidome (**Figure 4A**), with only BM CEs having low but significant inverse correlation with phospholipid classes in serum (O-PC, PE, SM). In a partial correlation network analysis, PFAS showed association with BM TGs, saturated FAs and BM LPCs (**Supplementary Figure 3**).

**Figure 4.**
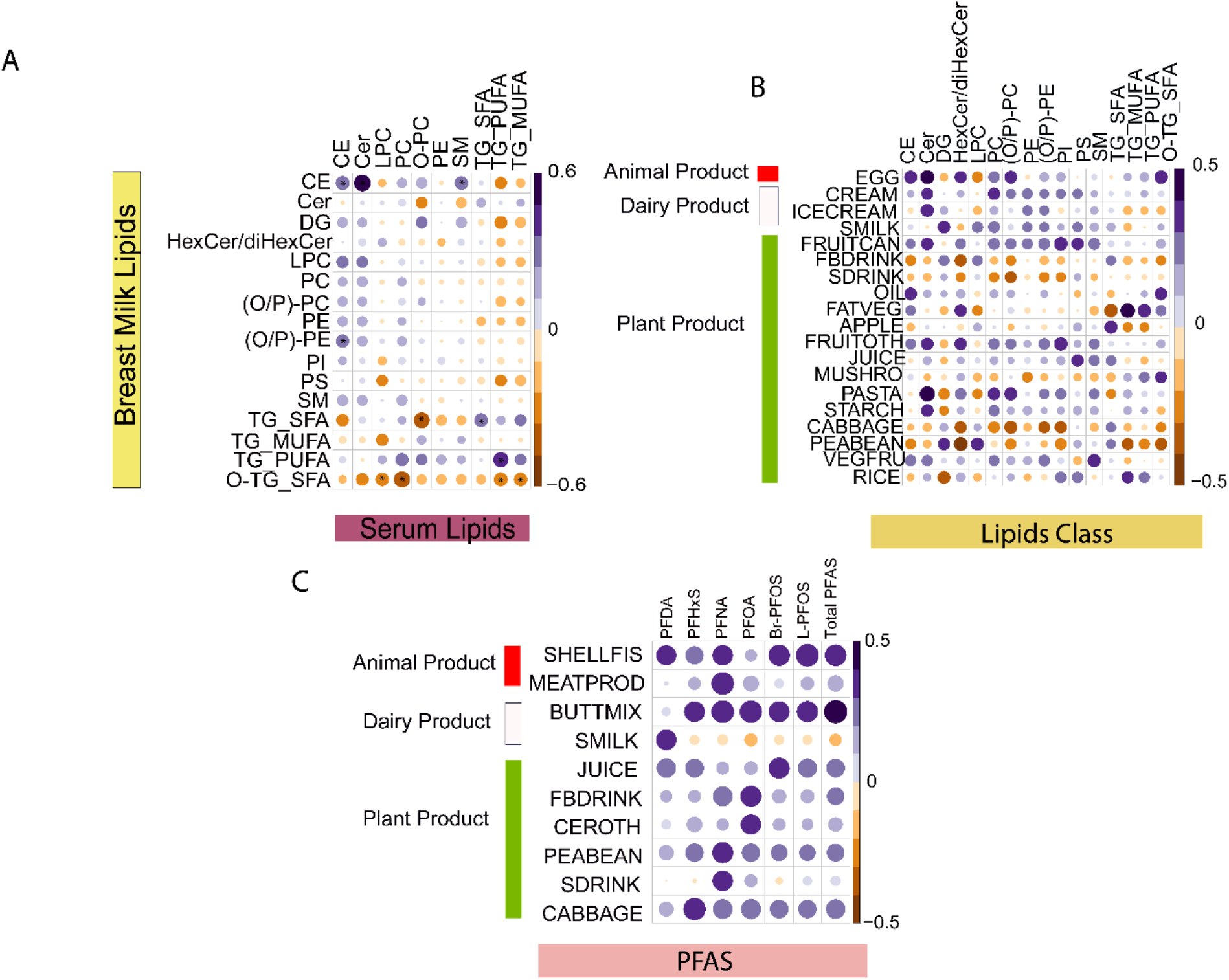
Associations of BM milk and serum lipids, PFAS exposure, and maternal diet. (**A**) Pairwise Spearman correlation between BM lipids and serum lipids at 3 months of age. (**B**) Correlation between BM lipids and dietary factors during pregnancy. (**C**) Correlation of PFAS exposure at 3 months of age and dietary factors during pregnancy. Positive correlations in purple, inverse correlations in orange Dot size for each pairwise correlation corresponds to the strength of the calculated correlation. *p < 0.05. Abbreviations: BUTTMIX, butter fat blends; CEROTH, other cereals (other than wheat, barley, rye, oats, rice); FATVEG, margarine and fat spreads >55%; FRUITOTH, other fruit and berries; MEATPROD, meat products; MUSHRO, mushroom; SDRINK and FBDDRINK, beverages; SHELFIS, shellfish; SMILK, sour milk products; VEGFRU vegetables.

The diet and PFAS levels during pregnancy were also assessed in relation to the BM lipidome (**Figure 4B-C**). The diet did not have a marked impact on the BM composition, with the most pronounced associations observed for egg and milk products, vegetable fat as well as some vegetable/fruit products. The partial correlation analysis also identified associations with specific lipid classes and cheese (DG), soya (Cer), rye (LPC), dietary fat (CE) and vegetables (PE). Stronger associations were observed between the diet and serum lipids. Also PFAS showed association with several dietary classes, including shellfish, meat products and some drinks (**Figure 4C**).

## Discussion

We found that the total lipids were down-regulated and the overall lipid composition was altered in mothers highly exposed to PFAS. Specifically, BM lipids containing saturated and monounsaturated fatty acids were increased, while lipids containing polyunsaturated fatty acyls were decreased in the highest exposure group at the beginning of breastfeeding. The BM samples collected at 3-month time point showed less association with the PFAS levels. Our results are in agreement with two previous studies, showing that exposure to BPA modifies milk yield and modified the synthesis and/or secretion of milk fat globules during early lactation in rats (*16,27*). The BPA exposure changed both the amount of total lipids and the FA composition in milk in dose dependent manner. The study also demonstrated that the mammary gland histo-architecture in the BPA-exposed F1 dams remained modified during early lactation (LD2), but not during mid-lactation (*27*), thus suggesting that the impact was more pronounced in the early lactation. Similar changes in mammary gland structure has been reported in PFAS-exposed animal models (*9, 10*).

The mechanisms controlling lactation are complex, involving several hormonal triggers such as peptide hormones (prolactin, oxytocin, vasopressin, and adrenocorticotropic hormone) and steroid hormones (glucocorticoids, estrogens, progestogens, and androgens). Importantly, PFOS is known to have weak estrogenic activities, whereas it exerts an antiestrogenic effect when co-administered with estradiol (*28, 29*). PFOS has also been reported to decrease production of estradiol and prolactin in women of reproductive age (*29, 30*) and to alter steroidogenesis (*31*). Indeed, several observational studies have shown that PFAS exposure is linked with several aspects of the reproductive system in females, such as delayed menarche, disruption of menstrual cycle regularity, early menopause and premature ovarian insufficiency and altered levels of circulating sex steroid hormones, as recently reviewed (*32*). The reduction of the total lipids as well as changes in the fatty acid composition of the lipids at high PFAS exposure, as observed in our study, could potentially suggest disturbed milk production and synthesis, possible due to disturbed interplay of estrogen, progesterone and prolactin by PFAS exposure, in addition to PFAS-triggered changes in mammary gland structure as reported in animal studies. More detailed studies are however required in order to verify this hypothesis. Interestingly, the PFAS-induced changes on BM lipidome in colostrum related to phospholipid composition were similar to those reported in mothers who have delivered prematurely (*33*), *i*.*e*., higher levels of phospholipids were observed in preterm colostrum and milk. The differences in the amount of phospholipids and in fatty acyl distribution in phospholipid species between human milk and infant formulas can imply biologically significant differences for newborn infants (*34*).

The PFAS exposure, particularly PFOS and PFHxS, were positively associated with PE levels in BM. Moreover, the PC/PE ratio was decreased with high level of PFAS exposure, with the main drivers of this effect being PFOS and PFOA. These polar lipids are mainly located on the milk fat globule (MFG) membrane which is structured as an ordered triple layer of phospholipids, cholesterol, and proteins, with PE, PI, and PS located mainly in the inner layer while PC and SM in the outer layer of the membrane (*35*). The PC/PE ratio reflects the size of the MFGs, with a high PC/PE ratio favoring small MFGs (*36*). Mechanistic studies have further implicated that in secretion of MFG from mammary epithelial cells, increasing PE content enhances fusion between lipid droplets, and therefore increases lipid droplet size (*37*). Thus, we conclude that the PFAS exposure was linked with increased size of the MFGs. The size of the MFGs is highly relevant, as small MFGs have relatively higher content of membrane compared with large globules, and as reported in several human and animal studies, this membrane exerts diverse positive health effects (*38, 39*). It is also known that the rate at which MFGs are digested is related to their diameter, with small particles more efficiently digested (*40*).

Interestingly, two PFAS (PFDA and PFNA) were associated with increased levels of fecal calprotectin, a protein released from infiltrating neutrophils and mucosal macrophages during inflammatory condition, which has been suggested as a biomarker of gastrointestinal (GI) inflammation (*41*). Recent studies indicatd that early gut dysbiosis in infants induces enteric inflammation, demonstrated by increased expression of fecal calprotectin (*42*). Increased intestinal inflammation, reduced gut barrier function and resulting influx of proinflammatory compounds can lead to activation of islet-specific T cells triggering autoimmune diabetes (*43, 44*). Recent study in highly exposed adults showed an opposite trend than that we observed in the current study, *i*.*e*., an inverse association between PFAS and fecal calprotetin. However, they studied only three PFAS substances (PFHxS, PFOS and PFOA) (*45*).

We further investigated whether the BM lipid composition was associated with the growth in the offspring, since multiple studies have shown that the lipid composition of the BM is highly relevant for further growth and development of the infant. The PUFAs are crucial nutrients, especially DHA and arachidonic acid (AA), which are involved in growth, the immune system, vision, and cognitive and motor development. It has been reported that PUFAs in colostrum were inversely associated with the infant growth at 6 months of age (*46*). It has been suggested that DHA during pregnancy, lactation and early life may be associated with significant benefits in infant growth and development (*47*). Indeed, we observed that the DHA-containing lipids showed a trend of being positively associated with the offspring growth, although the change was not statistically significant. These lipids were on the other hand at lower levels in the milk of highly exposed mothers. This may explain the inverse trend for the association between PFAS and infant growth. Interestingly, PEs were characterized by a significant inverse association with weight at 3 and 6months. Also the PC/PE ratio, indicative of the MFG size, was positively associated with growth, supporting the data showing more efficient digestion of the smaller MFGs. Differential MFG size and composition are involved in modulating the immune response (properties of T cells) and has impact on structural development of the intestine as well as maturation of infant gut microbiota (*40*).

Also several other BM lipids that were associated with PFAS exposure had a negative effect on the weight development of the infants while a similar impact was also observed with PFOA. Previous studies have indeed implicated that the lipids in the BM are the largest source of energy and they are associated not only with growth but also in the formation of the nervous system including brain and spinal cord, participation in neurobehavioral development and in correct retinal growth and visual acuity (*48*).

Our results also identified association between growth and PFAS exposure, which is in agreement with recent studies reporting that prenatal PFAS exposure is associated with lower weight and BMI in infants between 5 and 12 months of age (*49, 50*). Several studies have suggested that growth recovery in PFAS-exposed children may, however, occur later in infancy or early childhood (*51*). Those studies did not investigate the impact of the BM lipidome on growth, although most studies have been adjusted for breast-feeding. However, the impact on growth *via* altered BM lipid composition due to the PFAS exposure has not been investigated previously, and it is conceivable that breastfeeding may act as a causal intermediate on a pathway linking prenatal PFAS to infant growth.

We acknowledge some limitations in our study. The first shortcoming is the small sample size, which did not allow us to systematically understand the impact of co-factors such as prior breastfeeding, duration of breastfeeding and its relationship with the PFAS exposure. However, for the first time as far as we are aware, this study reports impacts of PFAS exposures on the molecular lipid content of human BM. Thus, our results provide potentially clinically significant findings for understanding the downstream impact of PFAS exposure in the infant *via* the nursing mothers, which needs validation in larger cohorts. Next, the direct measurement of the size of MFGs is missing, thus it may be an inaccurate assessment of the impact of MFGs and its role in infant development. Notwithstanding this, the major building components of the MFGs are the membrane phospholipids and growing evidence suggests its compositional dynamics is related to the size of the MFG, which influence infant gut maturation.

In summary, our results indicate that the PFAS exposure decreases the nutritional quality markers of human milk, with reduced total lipid content, decreased levels of PUFAs, including DHA and AA containing lipids, while the saturated lipids were increased with the PFAS exposure levels. Moreover, the changes in phospholipid composition indicated that the size of the MFGs were increased in the highly exposed group. These lipids were further associated with infant growth, with a clear trend that the PFAS levels were changing the composition of those BM lipids that favor infant growth.

## Data Availability

The lipidomics datasets generated in this study were submitted to the Metabolomics Workbench repository (https://www.metabolomicsworkbench.org).

## Acknowledgments

This study was supported by the Swedish Research Council (grant no. 2016-05176 to T.H and M.O), Formas (grant no. 2019-00869 to T.H and M.O), and the Novo Nordisk Foundation (Grant no. NNF20OC0063971 to T.H. and M.O.). The EDIA study was supported by the National Institute of Diabetes and Digestive and Kidney Diseases (NIDDK), National Institutes of Health (No. 1DP3DK094338-01 to M.K.), the Academy of Finland Centre of Excellence in Molecular Systems Immunology and Physiology Research 2012-17, No. 250114 to M.K. and M.O.).

Further support was received by the Academy of Finland postdoctoral grant (No. 323171 to S.L.) and the Medical Research Funds, Tampere and Helsinki University Hospitals (to M.K.).

## Figure captions

**Supplementary Table 1.**
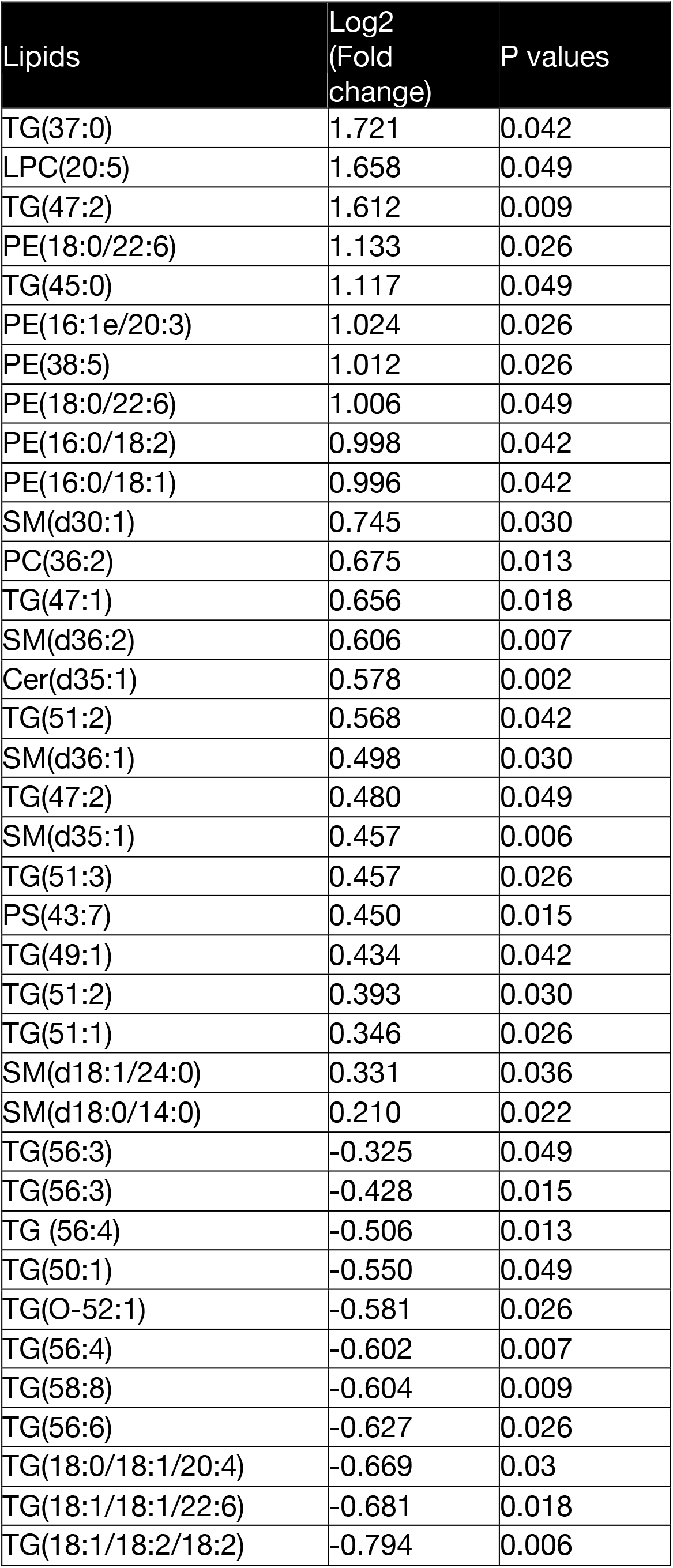
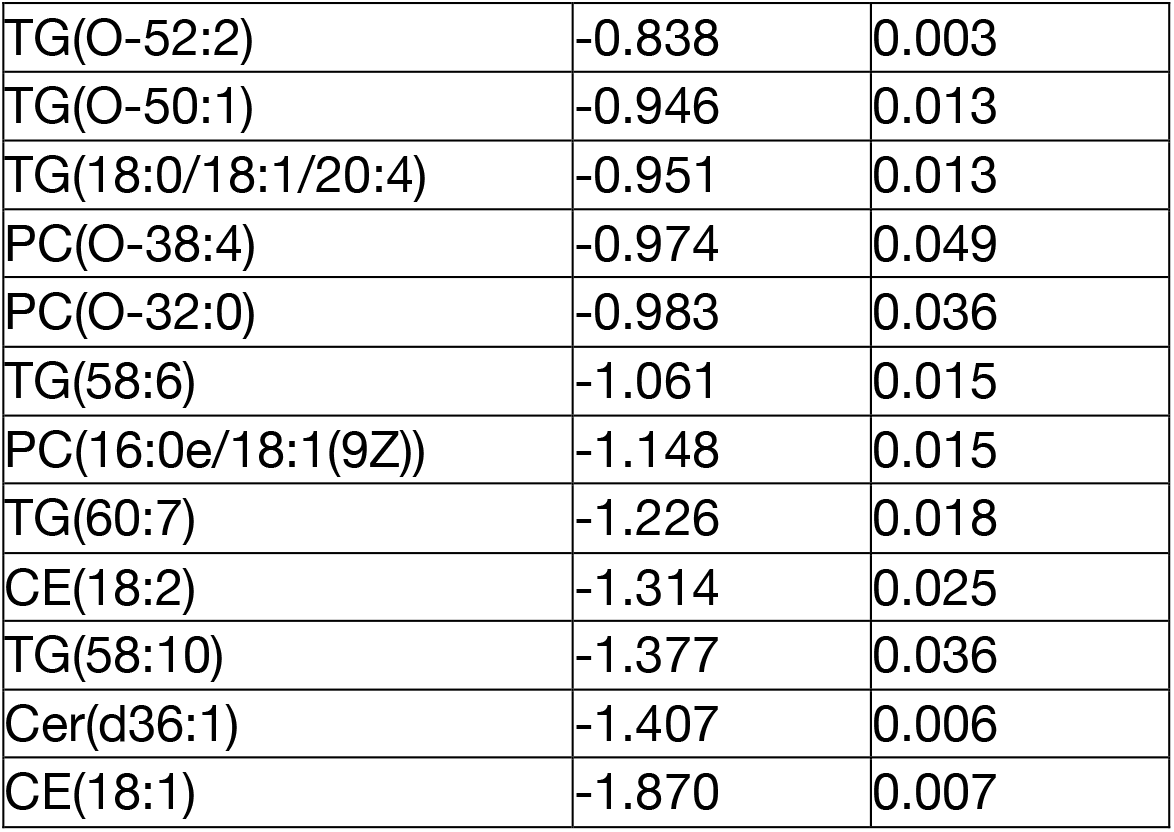
Differential breast milk lipids between high exposure (Q4) and low exposure (Q1) group at delivery.

**Supplementary Figure 1.**
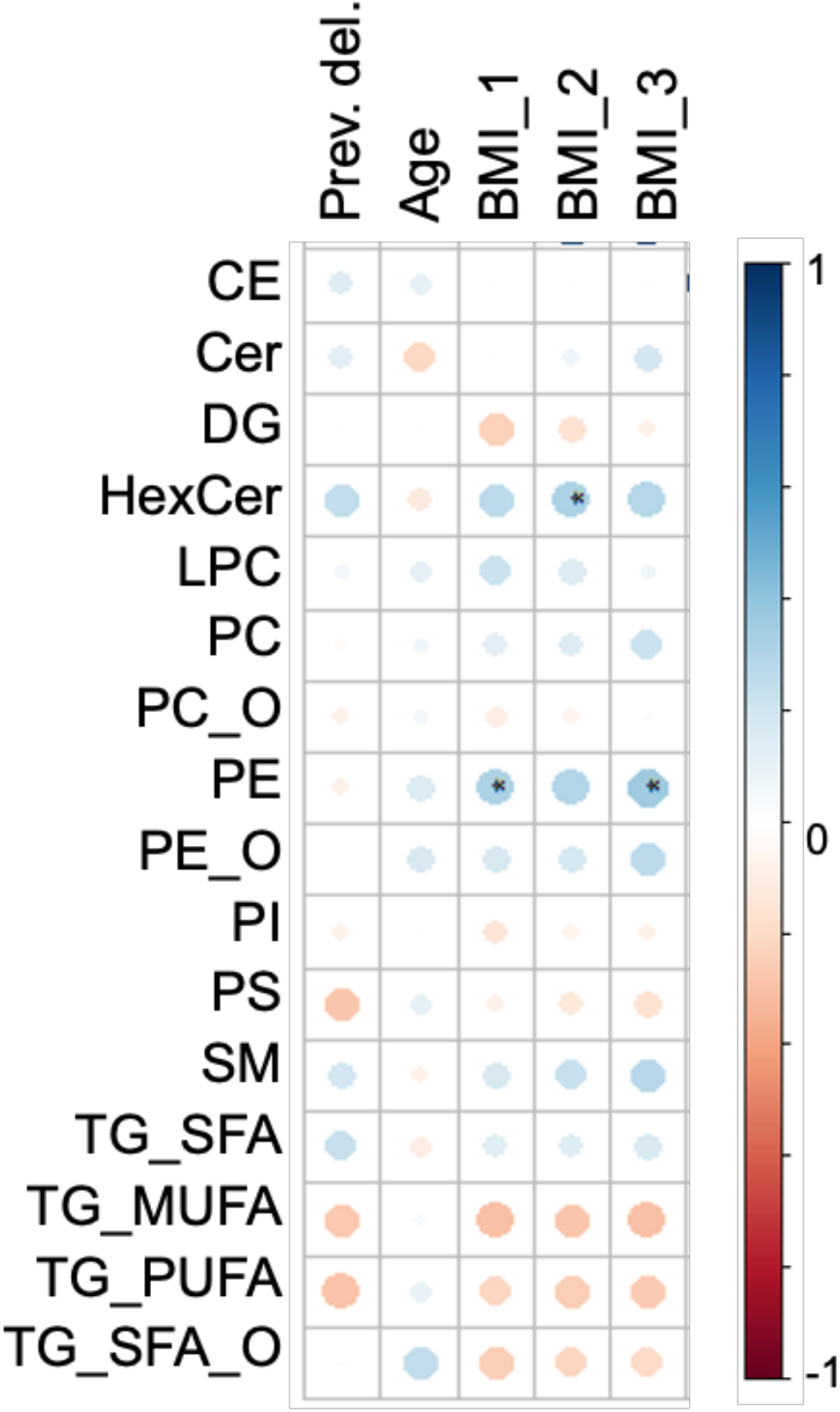
Correlation of BM lipids and demographic factors of the study participants. Positive correlations in blue, inverse correlations in red. Dot size for each pairwise correlation corresponds to the strength of the calculated correlation.

**Supplementary Figure 2.**
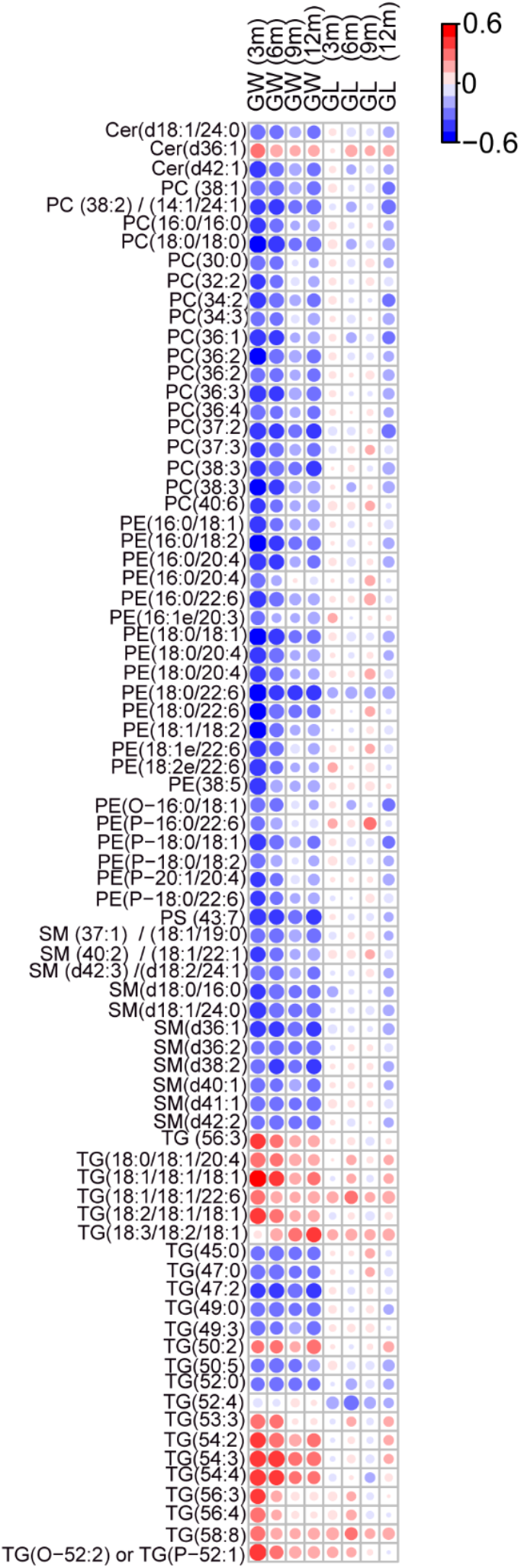
Correlation of individual breast milk lipids at delivery with offspring’s growth (weight or length) at age 3, 6, 9 and 12 months of age. Here, GW is growth in weight (kg/year), GL is growth in length (cm/year), FBD2 is Fecal beta defensin 2, FCP is fecal calprotectin. Correlations were calculated between simultaneous measurement at 3, 6, 9, and 12 months of age. GL (cm/year) and GW (kg/year) were calculated based on length in cm (or weight in kg) at given months – birth length in cm (or weight in kg))/time (measurement age (in years) at the given sampling day) respectively. Positive correlations in red, inverse correlations in blue. Dot size for each pairwise correlation corresponds to the strength of the calculated correlation.

**Supplementary Figure 3.**
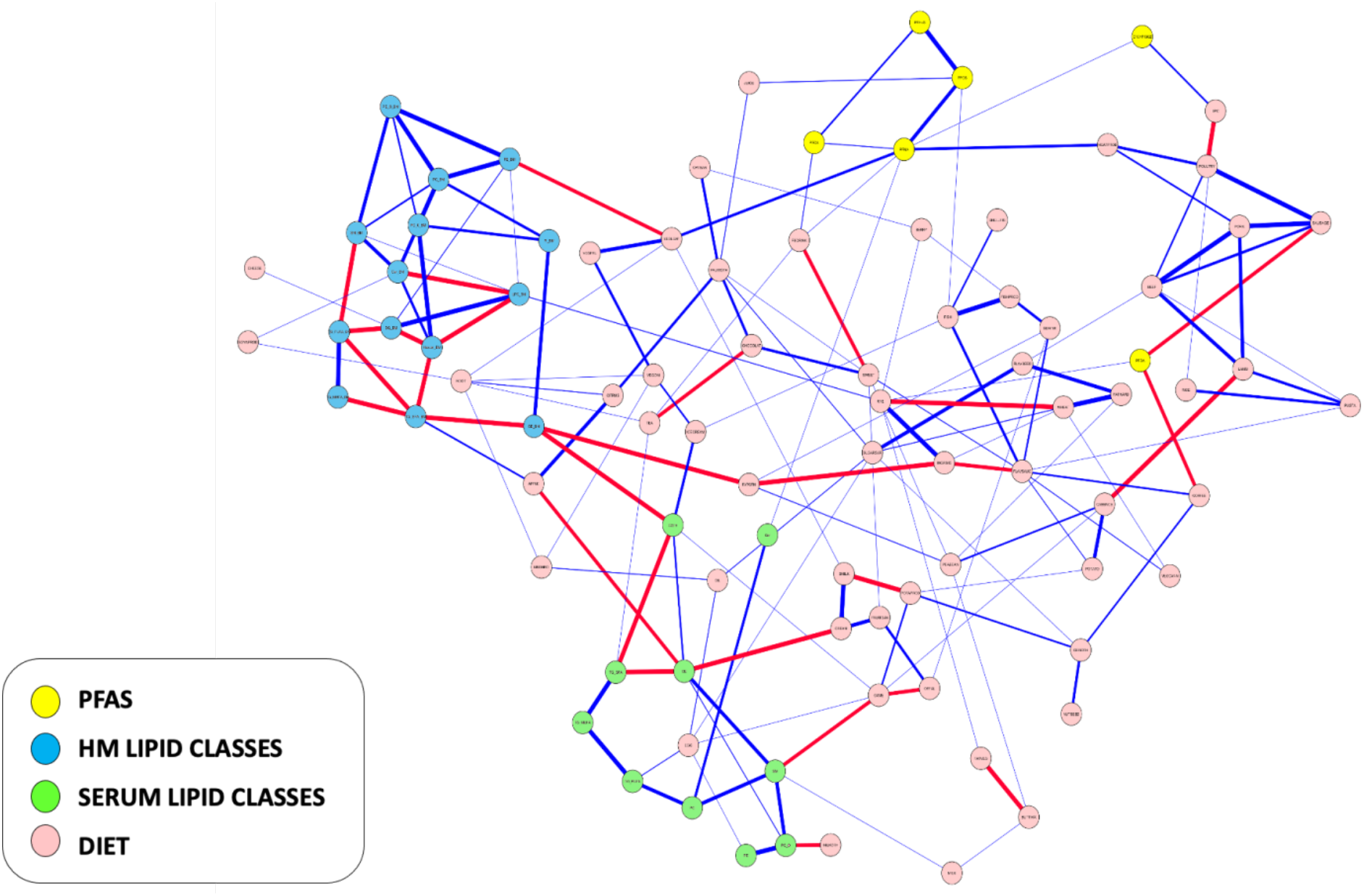
Partial correlation network analysis showing associations between maternal PFAS levels, serum lipidome, breast milk lipidome and diet during pregnancy.

## Notes

### Competing Interest Statement

The authors have declared no competing interest.

### Clinical Trial

NCT01735123

